# Acute Respiratory Distress Syndrome in Adults: A Retrospective Analysis of Temperature Trends, Demographic Factors, and Clinical Outcomes from the eICU Collaborative Research Database

**DOI:** 10.1101/2025.01.16.25320573

**Authors:** Ahmed Dawood Al Mahrizi, Charles Ezenwanne, Fatima Mossolem, Carlos Valladares, Harman Gill, Matthew Boyle

## Abstract

**Background:** Acute Respiratory Distress Syndrome is characterized by the sudden onset of hypoxemia, reduced lung compliance, and bilateral pulmonary infiltrates. We investigated patient demographics and clinical predictors of outcomes in adult patients with ARDS.

**Methods:** We conducted a retrospective multi-center intensive care unit database study using the eICU Collaborative Research Database (eICU-CRD). The Analysis included 304 patients with ARDS who met all inclusion criteria, including key demographic variables. We Assessed associations between these variables and outcomes, including mortality and length of stay (LoS).

**Results:** For each year of increased age, odds of mortality decreased by 4.7% (OR = 0.953, p < 0.001). Higher temperatures were associated with reduced mortality (OR=0.470, p=0.003). Gender (p = 0.596) and BMI (p = 0.964) were not significant predictors of mortality. A multiple linear regression model for predicting ICU length of stay was statistically significant at explaining the variance compared to the null model. High temperatures (β = 0.981, p = 0.015) and low temperatures (β = -1.038, p = 0.006) were significant predictors of LoS, while gender and BMI were not. There was significant association between ethnicity and hospital discharge status (χ^2^(5) = 13.123, p = 0.022), which suggests disparities in outcomes across ethnic groups.

**Conclusion:** Our study suggests that increasing age and higher temperatures may serve as protective factors, contrary to the popular belief of age-increasing fragility. Analysis also found that gender and BMI do not affect patient LoS nor mortality. These findings suggest clinicians should consider a patient’s age and core body temperature when assessing risk of mortality and prognosis, placing less emphasis on BMI and gender.

## Introduction

ARDS is an acute lung condition that causes low levels of oxygen, reduced lung compliance and pulmonary infiltrates. The disorder is characterized by endothelial cell damage, pulmonary edema, and diffuse alveolar damage [1]. Specifically, the disruption of the alveolar-capillary barrier leads to the leaking of proteins and fluid into the alveolar space, which leads to alveolar edema. This inflammation in the lungs creates widespread damage to both endothelial and epithelial cells that can lead to low blood oxygen and eventually a loss of functional lung tissue [2]. ARDS may result in respiratory failure and mechanical ventilation or death, possibly leading to death [3]. ARDS is both a localized lung condition and a systemic condition, with a high likelihood of impacting multiple organ systems [4]. The clinical presentation of ARDS typically develops within 72 hours after recognizing the underlying risk factor [2]. Presentation of ARDS is categorized by multiple symptoms including dyspnea, hypoxemia, tachypnea, cough, and chest discomfort [5]. Though there are multiple etiologies that incite ARDS, they share a common pathophysiologic mechanism that includes mechanical or infiltrative trauma impacting capillary and alveolar beds within the lung; this includes inflammation due to bacterial or viral pathogens [6, 7]. ARDS can occur due to direct lung injuries such as pneumonia, aspiration of gastric contents, or inhalation injuries. Indirect lung injuries, such as sepsis, trauma, pancreatitis, drug overdose, and transfusion-related acute lung injury, may also lead to ARDS [8]. The acute phase of lung injury is characterized by the body’s activation of its innate immune system and the activation of acute phase proteins [29]. Fever, defined as an increase in a person’s core body temperature greater than 0.5°C and serves to help eliminate infections from the body through mechanisms such as heat shock protein (HSP) activation. Tulapurkar et al. showed that the temperature increase occurring during fever is sufficient to alter endothelial and neutrophil function [39]. This then increases neutrophil transendothelial migration in vitro and recruitment in vivo [39].The diagnosis of ARDS is based on the Berlin definition, including a number of features regarding the timing of onset, chest imaging findings, origin of edema, and severity of hypoxemia according to the PaO2/FiO2 ratio [9, 3]. In particular, the American-European Consensus Conference defined ARDS as the acute onset of hypoxemia, an arterial pressure of oxygen to fraction of inspired oxygen (PaO2/FiO2) ≤ 200 mmHg, with bilateral infiltrate on frontal chest radiograph with no evidence of left atrial hypertension [10]. Furthermore, ARDS is classified into three categories: mild, moderate, and severe. Mild ARDS is diagnosed with a PaO2/FiO2 ratio between 200 and 300 mmHg. Moderate ARDS is a PaO2/FiO2 ratio between 100 and 200 mmHg. Whilst severe is a PaO2/FiO2 ratio below 100 mmHg [2].

Incidence of ARDS in the US ranges from 64 to 78 cases/100,000 person-years with 25% of cases classified as mild and 75% as moderate or severe [11]. Patient populations typically affected by ARDS are those with underlying health conditions such as chronic pulmonary disease or infection, cardiovascular disease, obesity, and heavy alcohol abuse [12]. A third of mild cases eventually progress to moderate or severe and 15% of all cases result in ICU admission [11]. ARDS accounts for approximately 10% of all ICU cases and carries a mortality rate of 40% in severe cases [13]. ARDS is typically treated with supportive care focused on increasing oxygen delivery, reducing acute inflammatory processes, and improving lung compliance. Increased oxygen delivery is often achieved with invasive airway ventilation using various techniques for pressure control and augmentation depending on the initial pulmonary insult and the clinical picture, since no pharmacological intervention has ever shown mortality benefit in these patients [14, 3]. Increasing lung compliance is achieved also through careful airway management as well as patient positioning [14].

There is growing appreciation in the literature for the multifactorial interactions affecting ARDS outcomes. High body temperature is related to a significantly lower risk of death [15]. Similarly, several studies have demonstrated that obese patients have lower mortality, though the mechanism of this relationship remains unclear [16]. Other reported determinants of poor outcomes in ARDS include increasing APACHE II scores, multiple organ failure hemodynamic instability, and elevated lactate levels [17]. In addition, there are certain risk factors that are linked to ARDS including age, chronic lung disease, obesity, and genetic factors. Older adults are more likely to contract ARDS due to having weakened immune systems, the presence of chronic conditions, and decreased lung compliance [18]. Conditions like chronic obstructive pulmonary disease (COPD) affect lung function and damage the tissue, which leads to an increased risk of ARDS [19]. In addition, studies have shown that obesity decreases lung compliance and worsens the mechanical ventilation process, which contributes to an increased risk of ARDS [20]. Previous studies have shown that specific polymorphisms like the -308G>A variant, are linked to increased susceptibility to ARDS [21].

This study will attempt to investigate patient demographic and clinical predictors of outcomes in a population of adult patients with ARDS, using data from the eICU Collaborative Research Database. Specific variables to be analyzed include age, sex, ethnicity, body mass index, temperature trends, and mortality in this critically ill population [13, 22, 15]. Identification of risk factors for poor outcomes will be useful for risk stratification and guiding management strategies in ARDS. By conducting a comprehensive analysis among a large and diverse population of patients with ARDS, this study aims to contribute to the expanding body of evidence on its prognosis, possibly offering new targets for therapeutic intervention.

## Methods

### Study Design and Data Source

We conducted a retrospective multi-center intensive care unit database study using Philips eICU collaborative research public database (eICU-CRD) [25, 26, 27]. This database was chosen for its solution of integration of different information systems and comprehensive methodology to handle diverse data types [23]. The eICU-CRD records and archives streaming data from teleICU monitoring, integrating information from multiple centers, making this database more comprehensive, reliable, and accessible for research [23, 25, 26, 27]. The database comprises health information of over 200,000 patients admitted to any one of the 335 ICUs at 208 hospitals located throughout the US from 2014-2015, and includes a comprehensive list of variables, like vital sign measurements, care plan documentation, severity of illness measures, diagnosis information, and treatment information [23]. The use of the database has been authorized by the review boards of the Massachusetts Institute of Technology (Cambridge, MA, USA). One of the authors, Ahmed D. Al Mahrizi has participated in a series of courses and obtained authorization to access the eICU-CRD after passing the required assessments (Certificate ID: 67053173), and was responsible for data extraction. All procedures conducted were within the regulations and guidelines of eICU-CRD.

### Study Population

The eICU-CRD captures continuous multi-center information on physiological measurements and clinical data in 200,000 ICU patients, enabling multiple patients with the same diagnosis to be studied simultaneously. This is particularly valuable in standardizing and classifying conditions such as Acute Respiratory Distress Syndrome (ARDS), which previously have been challenging due to the unstandardized diagnostic and therapeutic strategies associated with this syndrome [28]. We limited our patient population to patients diagnosed with ARDS, as indicated in the clinical data segment of the database, and filtered the patient records to analyze variables like gender, admission body mass index (Admission BMI), temperature extremes (highTempC_4, lowTempC_4), hospital discharge status, ICU length of stay (actualiculos), and ethnicity. We excluded records noting an “unknown” or “other” classification of gender or missing values, particularly in the variables of gender, admission BMI, temperature extremes, discharge status, and ICU length of stay (Supplement 1). The final analysis included 304 patients with acute respiratory distress syndrome who met all the other inclusion criteria boasting diverse ethnic backgrounds.

### Variables

The primary variables of interest were gender, admission BMI, temperature extremes (highTempC_4, lowTempC_4), hospital discharge status, ICU length of stay (actualiculos), and ethnicity.

### Statistical Analysis

Descriptive statistics and boxplots were generated for continuous variables (temperature extremes, age, length of stay), stratified by gender and ethnicity. Between-gender comparisons of continuous variables (ICU length of stay, age, admission BMI, temperature extremes) were conducted using independent samples t-tests, with effect sizes calculated as Cohen’s d using pooled standard deviation. All analyses used 95% confidence intervals. Associations between categorical variables were assessed using chi-squared tests. Pearson correlation coefficients quantified relationships between continuous variables. A logistic regression model with deviance residuals was developed to predict hospital discharge status, incorporating age, gender, BMI, temperature, length of stay, and ethnicity as predictors. A separate multiple linear regression model was constructed to predict the length of stay. K-means clustering was performed to identify patient subgroups based on clinical features. Clustering algorithm selection (k-means, median, or medioid) was determined by partitioning outcomes. Contingency tables were generated to depict the frequency distribution of categorical variables (ethnicity, gender, hospital discharge status) at once. Statistical significance was set at p < 0.05 for all analyses. All statistical analyses were conducted using JASP (Version 0.19.1) [24].

### Ethical Considerations

This study used a public database through the Philips eICU-CRD, which is exempt from informed consent, and has been deidentified by the MIT Laboratory for Computational Physiology, as approved by the institutional review boards of the Massachusetts Institute of Technology (No. 0403000206) and Beth Israel Deaconess Medical Center (2001-P-001699/14).

## Results

### Cohort Selection and Characteristics

During the one-year period (2014-2015), 200329 ICU patients were identified from the eICU Collaborative Research Database (eICU-CRD). 199524 patients were excluded on the basis of them not being diagnosed with ARDS in the clinical data segment of the database, 501 of which were excluded for having missing data on either gender, admission BMI, high temperature, low temperature, hospital discharge status, and actual ICU length of stay. Although it was a part of the exclusion criteria, no patients were excluded based on having a gender of “Other” or “Unknown.” 304 adult ICU patients with ARDS were analyzed (144 females, 160 males). The sample comprised various ethnic groups: Caucasians (n=222), African Americans (n=53), Other/Unknown (n=12), Hispanics (n=8), Asians (n=8), and Native Americans (n=1).

### Temperature Differences by Gender and Ethnicity

***Table 1*** shows descriptive statistics revealing a slight gender and racial difference in temperature measurements. Males exhibited marginally higher mean high temperatures (37.855°C, SD = 4.938) compared to females (37.391°C, SD = 0.605) ***(Figure 1)***. A similar pattern was observed for low temperatures (males: 36.625°C, SD = 5.201; females: 36.259°C, SD = 1.381). However, these differences were not statistically significant (high temperature: t(302) = -1.119, p = 0.264, 95% CI [-0.354, 0.097]; low temperature: t(302) = -0.820, p = 0.413, 95% CI [-0.319, 0.131]).

**Table 1:** Descriptive statistics model of high temperature, low temperature, age, and length of stay split by gender.

| Descriptive Statistics |  |  |  |  |  |  |  |  |
| --- | --- | --- | --- | --- | --- | --- | --- | --- |
|  | High Temperature |  | Low Temperature |  | Age |  | Length of Stay |  |
|  | Female | Male | Female | Male | Female | Male | Female | Male |
| Valid | 144 | 160 | 144 | 160 | 144 | 160 | 144 | 160 |
| Missing | 0 | 0 | 0 | 0 | 0 | 0 | 0 | 0 |
| Mean | 37.391 | 37.855 | 36.259 | 36.625 | 60.229 | 59.188 | 9.603 | 10.769 |
| Std. Deviation | 0.605 | 4.938 | 1.381 | 5.201 | 16.378 | 15.597 | 11.157 | 8.05 |
| Minimum | 36.2 | 35.836 | 21.1 | 17 | 22 | 18 | 0.364 | 0.896 |
| Maximum | 39.6 | 99.2 | 37.7 | 98.5 | 90 | 89 | 112.565 | 39.029 |

**Figure 1:**
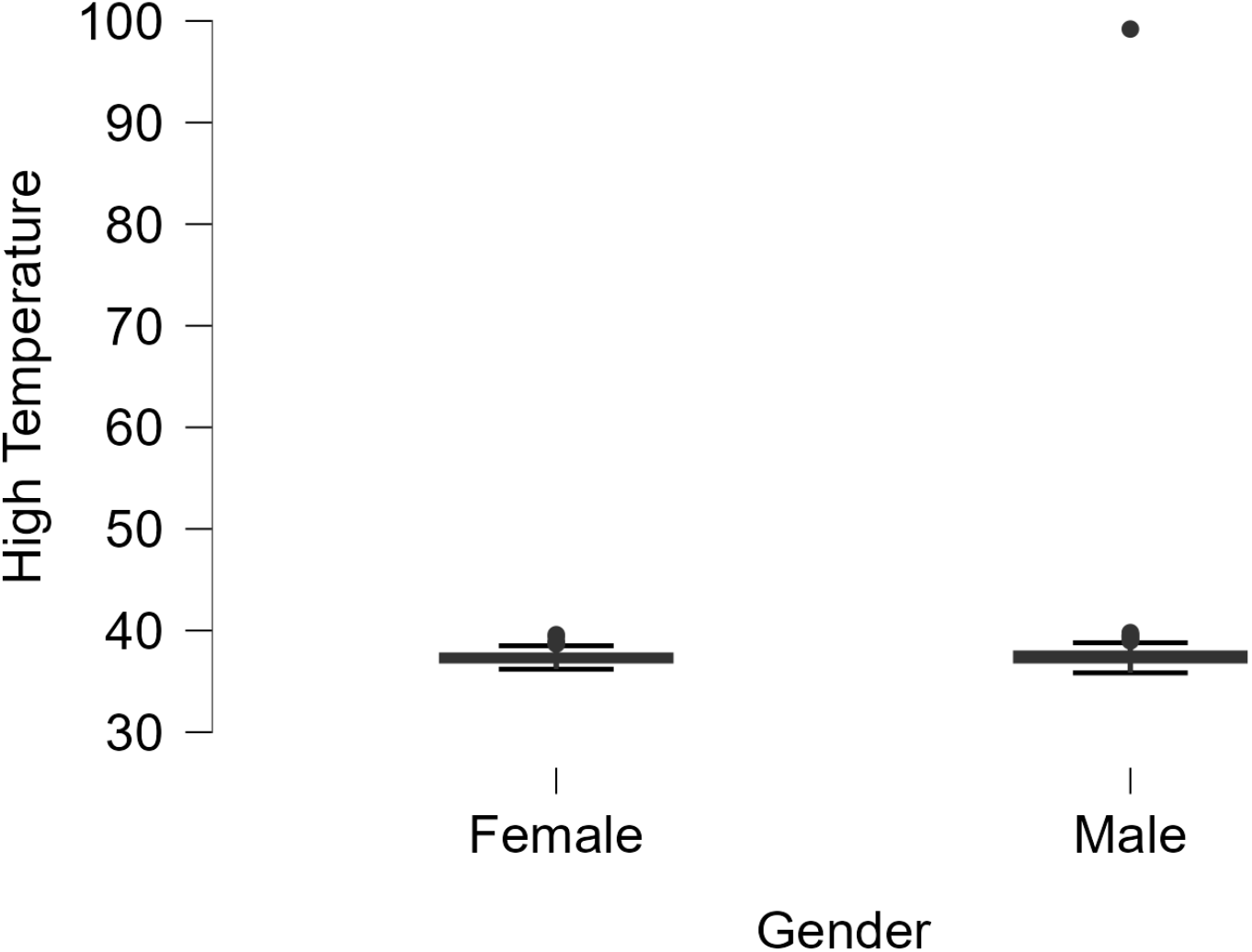
High Temperature boxplot split by patient gender

Across ethnic groups, Asians showed the highest mean high temperature (38.038°C), while African Americans had the lowest (37.346°C). For low temperatures, Hispanics had a notably lower mean (33.638°C) compared to other groups, as can be seen in ***Figure 2***.

**Figure 2:**
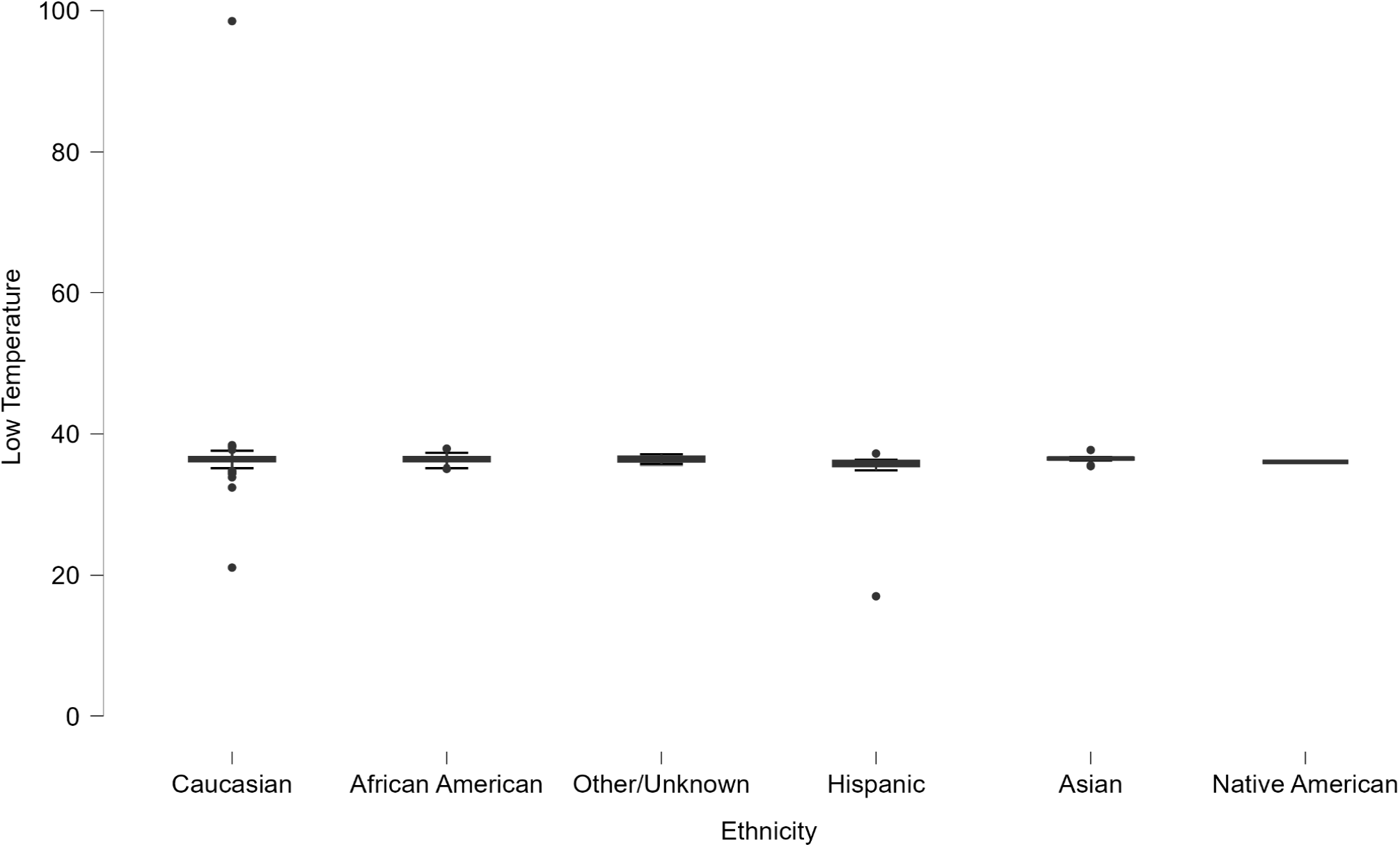
Low temperature boxplot split by patient ethnicity

### Predictors of Hospital Discharge Status

BMI data indicated that obesity (BMI ≥30) was present in 49% of patients overall. A logistic regression model predicting hospital discharge status yielded significant results for age (OR = 0.953, 95% CI [0.931, 0.975], p < 0.001) and high temperature (OR = 0.470, 95% CI [0.287, 0.770], p = 0.003). For each year of increase in age, the odds of mortality decreased by 4.7%, while higher temperatures were associated with lower mortality risk. Of note, gender (p = 0.596) and BMI (p = 0.964) were not significant predictors of mortality. The models demonstrated a moderate explanatory power (Nagelkerke R^2^ = 0.275).

### Predictors of Length of Stay

The multiple linear regression analysis shows that the overall model for predicting length of stay explains a statistically significant amount of variance compared to a null model (F(11, 292) = 2.875, p = 0.001, R^2^ = 0.098). High temperatures (β = 0.981, p = 0.015) and low temperatures (β = -1.038, p = 0.006) were significant positive predictors. However, gender and BMI were not significant predictors of length of stay ***(Figure 3)***.

**Figure 3:**
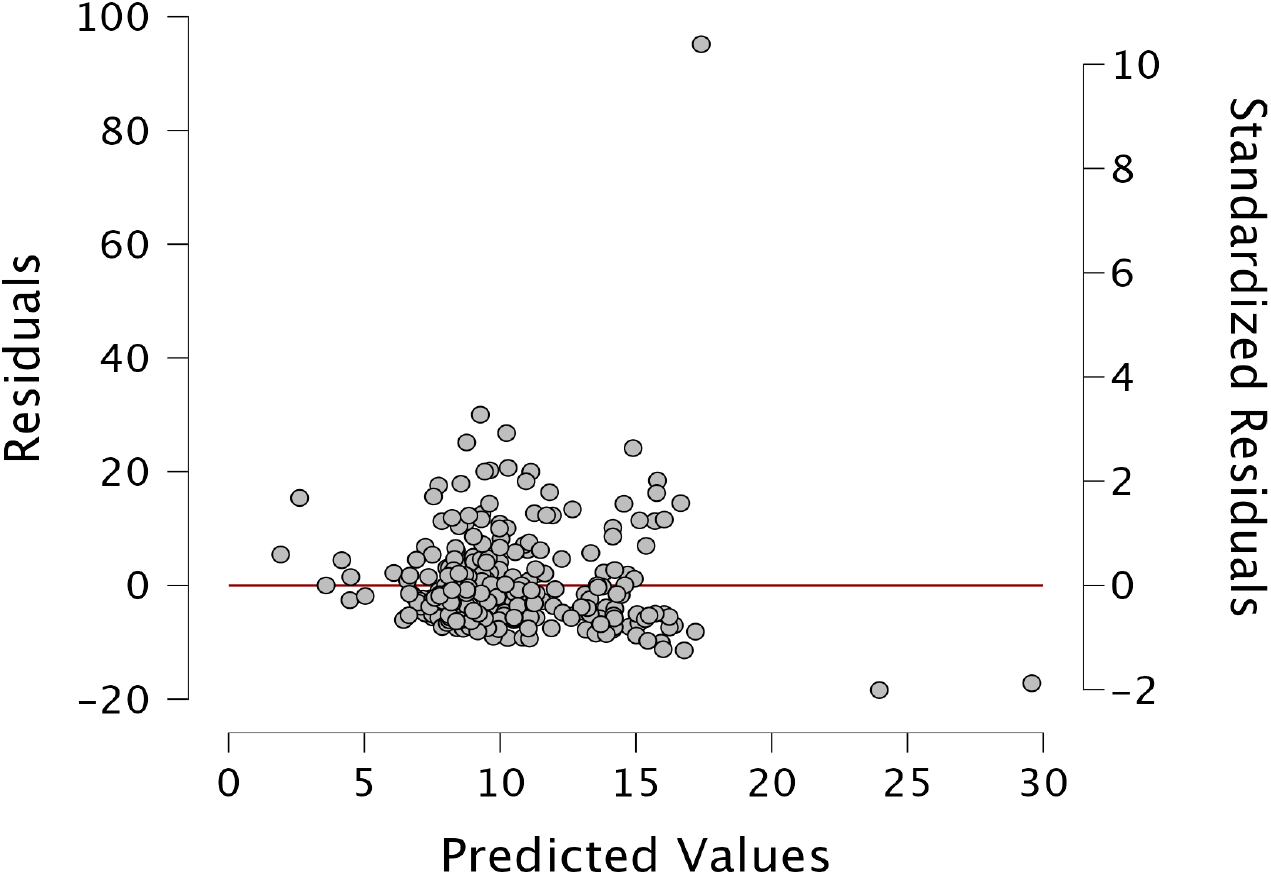
Residuals vs Predicted Plot for the linear regression model

### Ethnic Disparities in Outcomes

Chi-squared tests, as seen in ***Table 2***, revealed a significant association between ethnicity and hospital discharge status (χ^2^(5) = 13.123, p = 0.022), which suggests potential disparities in outcomes across ethnic groups.

**Table 2:** Chi-Squared Tests showing an association between ethnicity and hospital discharge status.

| Chi-Squared Tests |  |  |  |  |  |  |
| --- | --- | --- | --- | --- | --- | --- |
|  |  | Value |  | df |  | p |
| X<sup>2</sup> |  | 13.123 |  | 5 |  | 0.022 |
| N |  | 304 |  |  |  |  |

## Discussion

Our study examined the data of 304 adult patients, 144 females and 160 males, admitted into the ICU with the diagnosis of ARDS whose data was captured by eICU-CRD. Factors including BMI, age, temperature, and ethnicity were analyzed to determine their influence on patient outcomes such as length of stay and mortality. Among our findings, neither gender nor BMI were found to significantly impact mortality rates. In contrast, age and temperature demonstrated protective effects, showing an inverse relationship with mortality. Specifically, higher temperatures were associated with reduced mortality, while lower mean temperatures were linked to worse outcomes, particularly among African American and Hispanic patients. Although these results provide important insights, the limited sample size and absence of data on confounding variables, such as comorbidities and disease severity, may have influenced the conclusions. Even with these limitations, our findings emphasize the complexity of ARDS and highlight the need for clinicians to tailor their evaluations and interventions to each individual patient rather than relying on generalized demographic predictors. This study contributes to further understanding ARDS by identifying potential protective factors.

### Temperature as a Protective Factor

Higher temperatures are associated with decreased mortality, which has been identified in previous studies [15]. Guihur et al. performed a study looking at COVID-19 deaths that resulted from ARDS and demonstrated that mild fever promotes natural accumulation of HSP70, a protein that serves as a protective factor against apoptosis and maintains cellular homeostasis [30-32]. Singleton et al. showed that the knockout of HSP70 led to increased ARDS severity and higher mortality from sepsis [33]. These findings emphasize the potential protective effects of fever in ARDS patients and encourage further studies aimed to examine HSP70 levels as a potential biomarker for improved outcomes. Our regression model supports these findings and showed higher mean temperatures were associated with better outcomes. Ethnic differences in temperature trends were also observed. Our analysis showed that Asians had the highest mean high temperature, while African Americans had the lowest mean High Temperature. Our analysis also showed that Hispanics had a notably lower mean low temperature. Despite this trend, ethnicity was not identified as an independent predictor of mortality or length of stay in the regression model suggesting that while high temperatures may be protective, other factors influencing these disparities validate the need for further investigation.

Temperature trends also varied by gender, with males showing higher averages for both high and low temperature extremes than their female counterparts. This variation can be due to numerous factors, including size of the patient, disease severity, different measurement sites, which were not specified by the database [34]. Interestingly, our analysis showed no statistically significant difference in mortality based on gender, aligning with prior studies [45].

### Ethnic Disparities

Many previous studies have concluded that racial minorities in the United States do in fact have worse outcomes when admitted to the ICU with ARDS [35]. Whether these outcomes have to do with the primary disease or hospital wide complications, it is important for us to emphasize that race and ethnicity play a large part in patient care. While our analysis showed that temperature could play a protective role in the outcomes of ARDS, many studies have shown the opposite to be true. Wang et al. conducted a retrospective study on ARDS patients also infected with COVID-19 and concluded that temperatures greater than ≥39.1°C were actually detrimental to patient care [36]. Further studies analyzing the range of temperatures that could be deemed protective vs damaging may be necessary to further conclude their protective mechanisms. However the protective effect of increased temperatures seen here should help guide clinical care and not immediately be considered a negative prognostic factor when patients in the ICU develop a fever.

### Length of Stay

Length of stay was a factor we included due to its prognostic power, as patients that typically have longer hospital stays tend to have worse outcomes regardless of admission status [37]. This is due to many factors, primarily the increased risk of hospital related complications, especially nosocomial infections [38]. Both high temperatures and low temperatures were significant predictors of length of stay. Our analysis found that increased length of stay was negatively associated with mortality, indicating worse outcomes for patients that have longer hospital stays. Our findings align with previously mentioned physiological mechanisms [39], where fever-range temperatures enhance endothelial and neutrophil function, leading to increased neutrophil migration and recruitment. This temperature-dependent immune response may help explain the protective effect of fever observed in ARDS patients. Together, these findings suggest that fever could play a beneficial role in the inflammatory response during critical illness [15]. Our analysis using linear regression showed that high temperature and low temperature were both significant predictors of length of stay, along with discharge status, which indicated “Alive” or “Expired” at time of discharge. Gender also seemed to play a significant role in length of stay in our study with males having a longer average stay than females.. McNicholas et al. conducted a prospective cohort study that concluded, similarly, that women with ARDS had shorter lengths of stay [40]. It is important to note that while gender was seen to impact length of stay there was no significant difference in mortality between men and women within our study. Linear regression analysis also concluded that age, BMI, and ethnicity were not significant predictors of length of stay.

When running a Chi-squared analysis we saw a marginally significant association between ethnicity and gender (p = 0.058), as well as there being a significant association between ethnicity and hospital discharge status (p = 0.022), meaning that while there was no direct correlation between ethnicity, mortality and length of stay, it may still be a significant factor to consider when discussing patient care.

### Age as a Predictor

Age and ARDS have been previously studied; other studies have also noted a disproportionate impact of ARDS on younger populations, and especially so when diagnosed with COVID-19 [41]. Age played a surprising role in patient outcomes. Female patients admitted with ARDS were older on average than their male counterparts (60.229 years vs 59.188 years) however there was no significant difference in mortality based on gender. A logistic regression analysis predicting mortality showed that age actually had a statistically significant (0.953, p < 0.001) negative correlation with mortality. This was of particular interest to our team as many previous studies had concluded the opposite. Killien et al. conducted a study that saw higher mortality with increased age in ARDS patients following trauma [42]. However some studies have concluded that there may be a non-linear relationship between age and mortality, with those older than 55 years old being at the highest risk [43, 44]. Brown et al. concluded that age was a poor prognostic factor for even the development of ARDS [45]. As the average age of people around the world continues to increase it is important for clinicians to continuously adapt to the challenge of caring for an increasing geriatric population [46]. The analysis of this study showed that age increasing risk of death in patients with ARDS is not always true and we urge physicians to consider other factors of the patient’s condition when determining treatment plans.

### BMI and Gender

Within the patients analyzed, 49% were considered obese (BMI ≥30). We have concluded through logistic regression analysis that BMI was not a significant predictor of mortality or length of stay. Lou et al. explored this relationship in further detail showing a U-shaped association between BMI and ARDS Where the higher and lower BMI values both have an increased risk of developing ARDS [47]. They further concluded that a BMI ≥35 was also slightly correlated with patient death and had some statistical significance [47]. There remains debate between the role of BMI and patient outcomes, however, our study showed that BMI did not significantly contribute to mortality or length of stay.

### Limitations

The sample size of this study was fairly small, 304 patients, from one hospital system, this small sample may limit the generalizability of these results. While the patient sample was diverse, factors such as previous patient history, trauma, and lifestyle (i.e. smoking, alcohol use etc.) were not considered in this analysis. We also did not have long term follow up data for our patient population to be analyzed so further complications may arise that were not included in our analysis.

## Conclusion

This study suggests that in ARDS, the patient’s age and higher temperatures may serve as protective factors, contrary to the popular belief of age-increasing fragility. However, it is essential to note that there may have been other significant confounding variables present in the younger populations or selection bias that may serve to explain these findings, such as the presence of comorbidities among the younger population analyzed and the severity of the disease onset. The analysis also found that gender and BMI do not affect patient outcomes. These findings suggest that clinicians can disregard any gender or BMI adjustments that are standardized within the current management of ARDS patients. This study presents a foundation for the future management of ARDS patients and should be replicated using more extensive databases with greater patient diversity to further support our findings.

## Acknowledgement

We acknowledge the researchers at the MIT Laboratory for Computational Physiology for publicly sharing the eICU database. We would also like to thank Seth Spicer for his valuable assistance in coordinating the author group and facilitating communication among contributors and Kyle Copenhaver for his quality assurance support.

## Data Availability Statement

The data analyzed in this study was obtained from the eICU Collaborative Research Database (eICU-CRD), the following licenses/restrictions apply: To access the files, you must be a credentialed user, complete the required training (CITI Data or Specimens Only Research) and sign the user agreement for the project. Requests to access these datasets should be directed to PhysioNet, https://doi.org/10.13026/C2WM1R.

