## Supplement 1 for "Acute Respiratory Distress Syndrome in Adults: A Retrospective Analysis of Temperature Trends, Demographic Factors, and Clinical Outcomes from the eICU Collaborative Research Database"

**R Filter**

```
grepl("ARDS", apacheadmissiondx) &
!is.na(gender) &
!is.na(admissionBMI) &
!is.na(highTempC_4) &
!is.na(lowTempC_4) &
!is.na(hospitaldischargestatus) &
!is.na(actualiculos) &
hospitaldischargestatus != "Unknown" &
gender != "Unknown" &
gender != "Other"
```
