## Supplement 2 for "Acute Respiratory Distress Syndrome in Adults: A Retrospective Analysis of Temperature Trends, Demographic Factors, and Clinical Outcomes from the eICU Collaborative Research Database"

### Correlation Analysis of Clinical Parameters in ICU Patients with ARDS

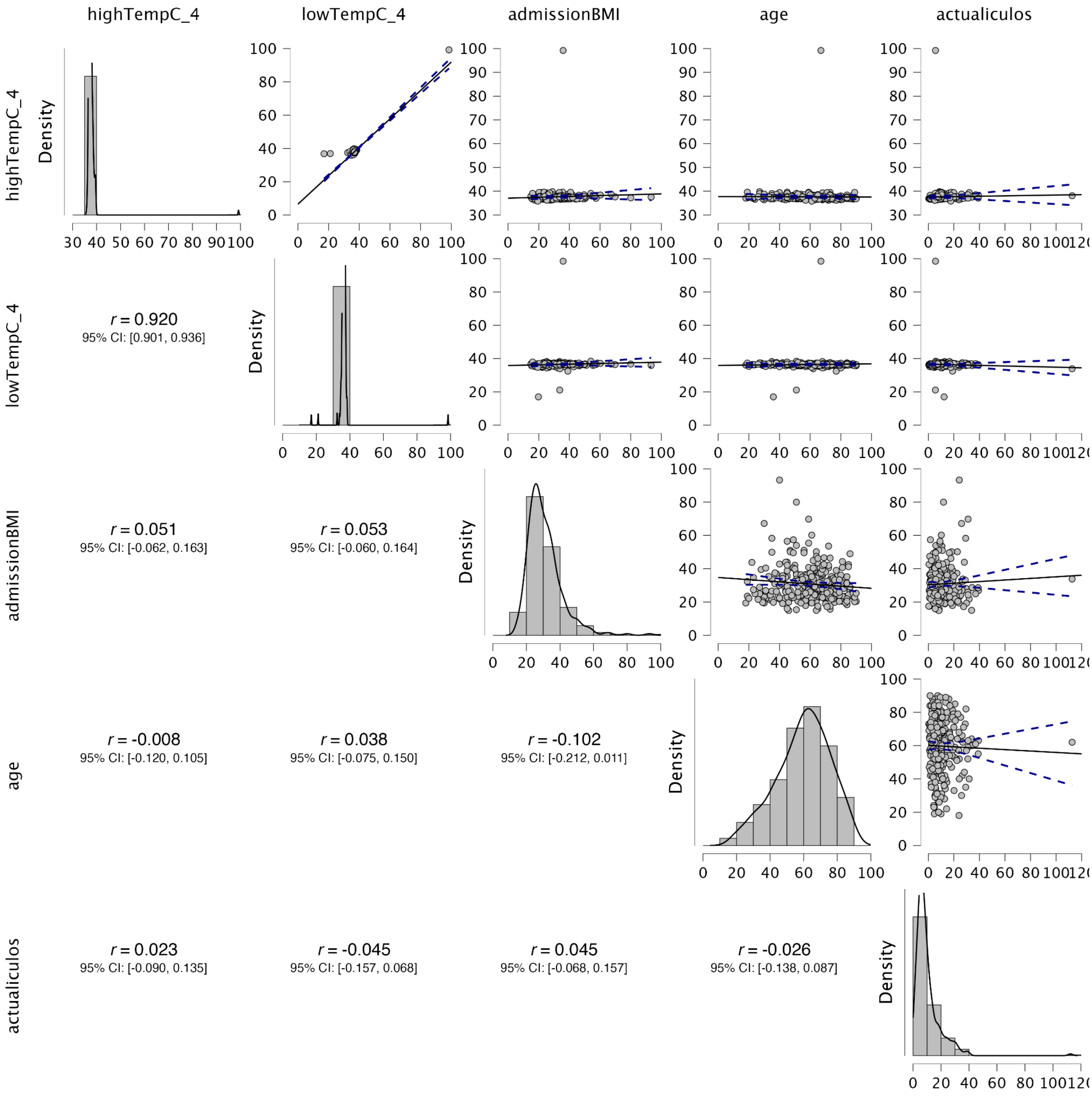


*(Figure 1: Correlation matrix displaying pairwise relationships between clinical parameters)*

(Fig. 1) Correlation analysis show age and admission BMI appear to have a slight negative correlation (r = -0.102), The distribution plots show that age follows a roughly normal distribution centered around middle age, while BMI has a right-skewed distribution. Lastly, ICU length of stay (actualiculos) is heavily right-skewed, indicating most patients have shorter stays while some have extended periods
