## Supplement 3 for "Acute Respiratory Distress Syndrome in Adults: A Retrospective Analysis of Temperature Trends, Demographic Factors, and Clinical Outcomes from the eICU Collaborative Research Database"

**Exploratory Cluster Analysis Suggests Potential ARDS Patient Subgroups**

K-means neighborhood-based clustering data shows subtle groupings of ARDS patients with clinically different measurements. *Table 3* The model identified 4 subgroup clusters and 1 single-patient cluster from 304 patients with an $R^2$ value of 0.596 suggesting moderate explanatory power.

*Model Summary: K-Means Clustering*

| Clusters | N | $R^2$ | AIC | BIC | Silhouette |
|---|---|---|---|---|---|
| 5 | 304 | 0.596 | 795.080 | 906.590 | 0.240 |

*Note.* The model is optimized with respect to the *silhouette* value.

*(Table S1: Model summary table showing summary of k-means clustering model)*

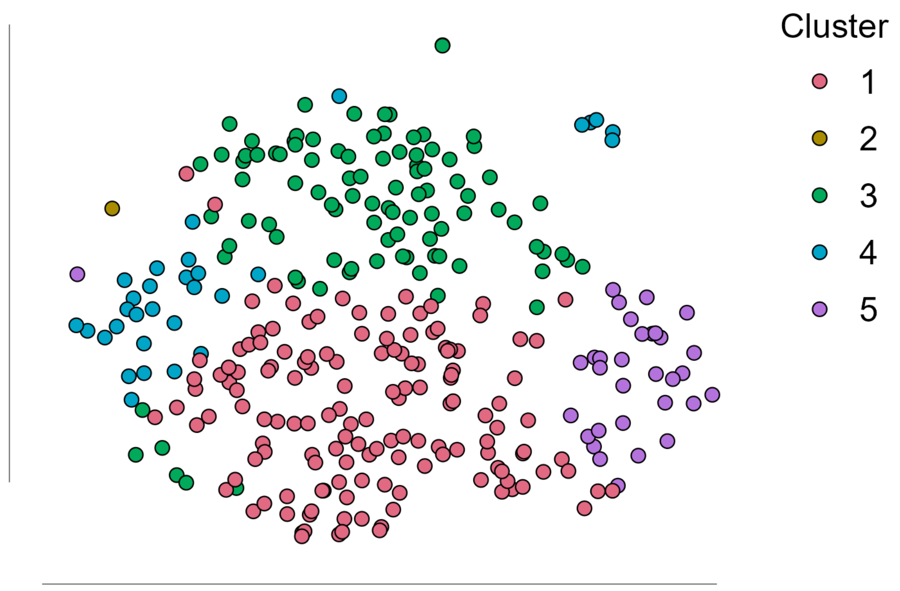

*(Figure S2 Two-dimensional t-SNE visualization of patient clusters showing distinct groupings of ARDS patients based on their clinical characteristics, where each point represents a patient and colors indicate cluster membership. )*

*Cluster Information*

| Cluster | 1 | 2 | 3 | 4 | 5 |
|---|---|---|---|---|---|
| Size | 143 | 1 | 97 | 31 | 32 |
| Explained proportion within-cluster heterogeneity | 0.316 | 0.000 | 0.304 | 0.153 | 0.228 |
| Within sum of squares | 232.038 | 0.000 | 223.143 | 112.383 | 167.520 |
| Silhouette score | 0.302 | 0.000 | 0.212 | 0.157 | 0.117 |
| Center age | 0.668 | 0.459 | -0.895 | -0.548 | 0.245 |
| Center admissionBMI | -0.197 | 0.494 | -0.288 | 2.084 | -0.280 |
| Center actualiculos | -0.296 | -0.476 | -0.237 | 0.144 | 1.915 |
| Center apachescore | 0.278 | 0.871 | -0.718 | -0.099 | 1.003 |
| Center highTempC_4 | -0.085 | 17.060 | -0.063 | 0.051 | -0.013 |
| Center lowTempC_4 | -0.020 | 15.953 | -0.115 | 0.037 | -0.097 |

*Note.* The Total Sum of Squares of the 5 cluster model is 1818

*(Table S2: Cluster information displaying the patient size of each cluster, the explained proportion of within-cluster heterogeneity. The latter is the cluster within the sum of squares divided by its total over the various clusters. It also displays the Centers (Standard Deviation from mean) of each feature)*

*(Figure 2)* Cluster 5 (32 patients) exhibit a subgroup of ARDS patients with notably longer ICU stays (SD actuallosicu: 1.915) and higher APACHE scores (SD apachescore: 1.003), suggesting a subgroup with more severe illness. Cluster 1, the largest group (143 patients), shows relatively average values across most measures, possibly representing a "typical" ARDS patient profile. Cluster 3 (97 patients) shows lower age scores (SD age: -0.895) and lower APACHE scores (SD -0.718), potentially representing a younger, less severely ill subgroup. Cluster 4 (31 patients) stands out for having higher BMI values (Center admissionBMI: 2.084), which might indicate a relationship between body mass and ARDS presentation.

### Limitations:

(Figure 2) The low Silhouette score (0.210) suggests overlap between clusters and lack of clustering separation. While there are subtle trends for these clusters, the within-cluster heterogeneity for some clusters (Cluster 1 and Cluster 3) are higher than expected >0.3 indicating there is some variation within these clusters. The clustering data can be used to explore more of the subphenotypes of ARDS.
