## Supplement 4 for "Acute Respiratory Distress Syndrome in Adults: A Retrospective Analysis of Temperature Trends, Demographic Factors, and Clinical Outcomes from the eICU Collaborative Research Database"

**ARDS Prevalence across Ethnicity and Gender**

**Contingency Tables**

| | | gender | | |
|---|---|---|---|---|
| **ethnicity** | | **Female** | **Male** | **Total** |
| Caucasian | Count | 100 | 122 | 222 |
| | % within row | 45.045 % | 54.955 % | 100.000 % |
| | % within column | 69.444 % | 76.250 % | 73.026 % |
| | % of total | 32.895 % | 40.132 % | 73.026 % |
| African American | Count | 24 | 29 | 53 |
| | % within row | 45.283 % | 54.717 % | 100.000 % |
| | % within column | 16.667 % | 18.125 % | 17.434 % |
| | % of total | 7.895 % | 9.539 % | 17.434 % |
| Other/Unknown | Count | 10 | 2 | 12 |
| | % within row | 83.333 % | 16.667 % | 100.000 % |
| | % within column | 6.944 % | 1.250 % | 3.947 % |
| | % of total | 3.289 % | 0.658 % | 3.947 % |
| Hispanic | Count | 3 | 5 | 8 |
| | % within row | 37.500 % | 62.500 % | 100.000 % |
| | % within column | 2.083 % | 3.125 % | 2.632 % |
| | % of total | 0.987 % | 1.645 % | 2.632 % |
| Asian | Count | 6 | 2 | 8 |
| | % within row | 75.000 % | 25.000 % | 100.000 % |
| | % within column | 4.167 % | 1.250 % | 2.632 % |
| | % of total | 1.974 % | 0.658 % | 2.632 % |
| Native American | Count | 1 | 0 | 1 |
| | % within row | 100.000 % | 0.000 % | 100.000 % |
| | % within column | 0.694 % | 0.000 % | 0.329 % |
| | % of total | 0.329 % | 0.000 % | 0.329 % |
| Total | Count | 144 | 160 | 304 |
| | % within row | 47.368 % | 52.632 % | 100.000 % |
| | % within column | 100.000 % | 100.000 % | 100.000 % |

| | | | | |
|---|---|---|---|---|
| | % of total | 47.368 % | 52.632 % | 100.000 % |

**Chi-Squared Tests**

| | Value | df | p |
|---|---|---|---|
| $X^2$ | 10.673 | 5 | 0.058 |
| N | 304 | | |

**Nominal**

| | Value |
|---|---|
| Contingency coefficient | 0.203 |
| Phi-coefficient | NaN |
| Cramer's V | 0.208 |
| Lambda (rows) | 0 |
| Lambda (columns) | 0 |
| Lambda (symmetric) | 0 |

(Table S3: Contingency Table represents the relationships between categorical variables (Ethnicity, Gender) and statistical tests are performed)

This contingency table analyzes the distribution of ARDS (Acute Respiratory Distress Syndrome) patients across ethnicity and gender. Of the sample Demographics (N=304), 47.37% were female and 52.63% were male. The ethnic distribution was Caucasian: 73.03% (222 patients) which made up the largest group, African American: 17.43% (53 patients) and other groups: each <4% of sample. Statistical Analysis: Chi-square test ($\chi^2 = 10.673$, df = 5, p = 0.058) suggests a marginally non-significant relationship between ethnicity and gender, Weak association strength (Cramer's V = 0.208, Contingency coefficient = 0.203) and Lambda values of 0 indicate poor predictive ability between variables
