## Supplement 5 for "Acute Respiratory Distress Syndrome in Adults: A Retrospective Analysis of Temperature Trends, Demographic Factors, and Clinical Outcomes from the eICU Collaborative Research Database"

### ARDS Discharge outcomes across Ethnicity

| *Contingency Tables* | | | | |
|---|---|---|---|---|
| | | hospital discharge status | | |
| ethnicity | | Expired | Alive | Total |
| Caucasian | Count | 46 | 176 | 222 |
| | % within row | 20.721 % | 79.279 % | 100.000 % |
| | % within column | 77.966 % | 71.837 % | 73.026 % |
| | % of total | 15.132 % | 57.895 % | 73.026 % |
| African American | Count | 5 | 48 | 53 |
| | % within row | 9.434 % | 90.566 % | 100.000 % |
| | % within column | 8.475 % | 19.592 % | 17.434 % |
| | % of total | 1.645 % | 15.789 % | 17.434 % |
| Other/Unknown | Count | 6 | 6 | 12 |
| | % within row | 50.000 % | 50.000 % | 100.000 % |
| | % within column | 10.169 % | 2.449 % | 3.947 % |
| | % of total | 1.974 % | 1.974 % | 3.947 % |
| Hispanic | Count | 2 | 6 | 8 |
| | % within row | 25.000 % | 75.000 % | 100.000 % |
| | % within column | 3.390 % | 2.449 % | 2.632 % |
| | % of total | 0.658 % | 1.974 % | 2.632 % |
| Asian | Count | 0 | 8 | 8 |
| | % within row | 0.000 % | 100.000 % | 100.000 % |
| | % within column | 0.000 % | 3.265 % | 2.632 % |
| | % of total | 0.000 % | 2.632 % | 2.632 % |
| Native American | Count | 0 | 1 | 1 |
| | % within row | 0.000 % | 100.000 % | 100.000 % |
| | % within column | 0.000 % | 0.408 % | 0.329 % |
| | % of total | 0.000 % | 0.329 % | 0.329 % |
| Total | Count | 59 | 245 | 304 |
| | % within row | 19.408 % | 80.592 % | 100.000 % |
| | % within column | 100.000 % | 100.000 % | 100.000 % |
| | % of total | 19.408 % | 80.592 % | 100.000 % |

| Chi-Squared Tests | | | |
|---|---|---|---|
| | **Value** | **df** | **p** |
| $X^2$ | 13.123 | 5 | 0.022 |
| N | 304 | | |

| Nominal | |
|---|---|
| | **Value** |
| Contingency coefficient | 0.203 |
| Phi-coefficient | NaN |
| Cramer's V | 0.208 |
| Lambda (rows) | 0 |
| Lambda (columns) | 0 |
| Lambda (symmetric) | 0 |

(Table S4: Contingency Table represents the relationships between categorical variables: Hospital discharge status and statistical tests are performed)

This contingency table examines survival outcomes (Expired vs. Alive) across different ethnic groups. The overall survival rate in the cohort is favorable, with 80.59% of patients surviving (245/304) and 19.41% (59/304) expired. Caucasians represent the largest group (73.03%, 222 patients), with a 79.28% survival rate. African Americans, the second-largest group (17.43%, 53 patients), showed a survival rate of 90.57%.
