## Supplement 6 for "Acute Respiratory Distress Syndrome in Adults: A Retrospective Analysis of Temperature Trends, Demographic Factors, and Clinical Outcomes from the eICU Collaborative Research Database"

**Temperature Significantly Influences Patient Outcomes**

| Model Summary - actualiculos | | | | | | | | |
|---|---|---|---|---|---|---|---|---|
| Model | R | $R^2$ | Adjusted $R^2$ | RMSE | $R^2$ Change | df1 | df2 | p |
| $M_0$ | 0 | 0 | 0 | 9.649 | 0 | 0 | 303 | |
| $M_1$ | 0.313 | 0.098 | 0.064 | 9.336 | 0.098 | 11 | 292 | 0.001 |

| ANOVA | | | | | | |
|---|---|---|---|---|---|---|
| Model | | Sum of Squares | df | Mean Square | F | p |
| $M_1$ | Regression | 2756.566 | 11 | 250.597 | 2.875 | 0.001 |
| | Residual | 25451.632 | 292 | 87.163 | | |
| | Total | 28208.198 | 303 | | | |

| Coefficients | | | | | | | | |
|---|---|---|---|---|---|---|---|---|
| | | | | | | | 95% CI | |
| Model | | Unstandardized | Standard Error | Standardized[a] | t | p | Lower | Upper |
| $M_0$ | (Intercept) | 10.217 | 0.553 | | 18.462 | < .001 | 9.128 | 11.306 |
| $M_1$ | (Intercept) | 15.494 | 6.765 | | 2.29 | 0.023 | 2.179 | 28.809 |
| | highTempC_4 | 0.981 | 0.402 | 0.367 | 2.438 | 0.015 | 0.189 | 1.772 |
| | lowTempC_4 | -1.038 | 0.372 | -0.418 | -2.786 | 0.006 | -1.771 | -0.305 |
| | admissionBMI | 0.041 | 0.053 | 0.043 | 0.771 | 0.442 | -0.064 | 0.146 |
| | age | -0.027 | 0.035 | -0.044 | -0.759 | 0.449 | -0.096 | 0.043 |
| | gender (Male) | 1.092 | 1.095 | | 0.997 | 0.32 | -1.064 | 3.248 |
| | ethnicity (African American) | 1.729 | 1.448 | | 1.194 | 0.234 | -1.121 | 4.579 |
| | ethnicity (Other/Unk | -0.573 | 2.828 | | -0.203 | 0.84 | -6.139 | 4.993 |

| | | | | | | | | |
|---|---|---|---|---|---|---|---|---|
| | nown) | | | | | | | |
| | ethnicity (Hispanic) | -5.303 | 3.492 | | -1.519 | 0.13 | -12.175 | 1.569 |
| | ethnicity (Asian) | -0.595 | 3.404 | | -0.175 | 0.861 | -7.295 | 6.104 |
| | ethnicity (Native American) | -4.98 | 9.412 | | -0.529 | 0.597 | -23.505 | 13.544 |
| | hospitaldischargestatus (Alive) | -5.839 | 1.441 | | -4.052 | < .001 | -8.674 | -3.003 |

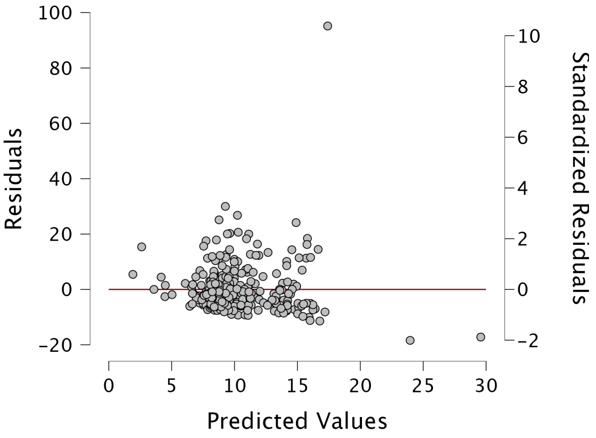


Residual vs predicted

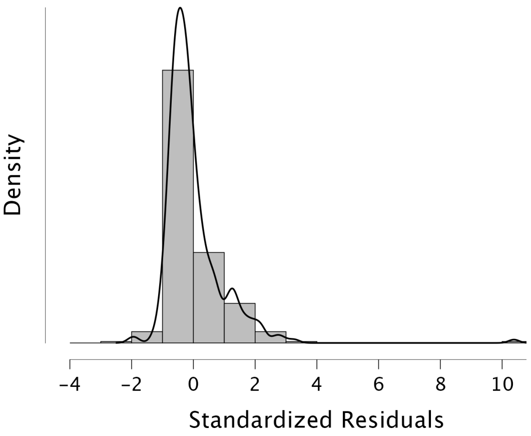


Standardized Residual Histogram

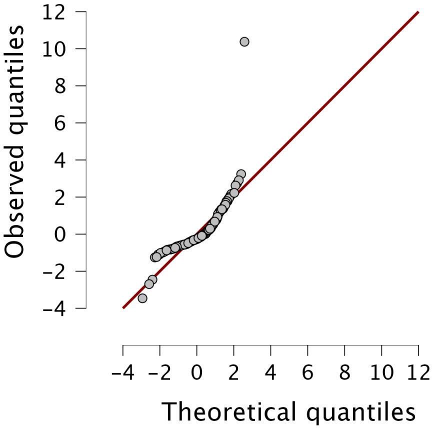


Q-Q Plot Standardized Residual

A regression analysis of hospital patient data reveals significant relationships between temperature conditions and patient outcomes. High temperatures were associated with increased outcome measures (β = 0.981, p = 0.015), while lower temperatures showed an inverse relationship of similar magnitude (β = -1.038, p = 0.006). These opposing effects of high and low temperatures suggest that temperature regulation may play a crucial role in patient care. The findings remained significant even after controlling for various demographic factors including age, BMI, gender, and ethnicity, none of which showed significant associations with the outcome.
